# Epidemiological investigation and intergenerational clinical characteristics of 24 COVID-19 patients associated with supermarket cluster

**DOI:** 10.1101/2020.04.11.20058891

**Authors:** Suochen Tian, Min Wu, Zhenqin Chang, Yunxia Wang, Guijie Zhou, Wenming Zhang, Junmin Xing, Hui Tian, Xihong Zhang, Xiuli Zou, Lina Zhang, Mingxin Liu, Juan Chen, Jian Han, Kang Ning, Shuangfeng Chen, Tiejun Wu

**Affiliations:** Department of Intensive Care Unit, Liaocheng people’s hospital, Liaocheng, Shandong, 252000, China; Department of Healthcare Associated Infection, Liaocheng people’s hospital, Liaocheng, Shandong, 252000, China; Department of Hemodialysis, Liaocheng people’s hospital, Liaocheng, Shandong, 252000, China; Department of Intensive Care Unit, Liaocheng Infectious Diseases Hospital, Liaocheng, Shandong, 252000, China; Intelligence Library Center, Liaocheng people’s hospital, Liaocheng, Shandong, 252000, China; Department of Respiratory Medicine, Liaocheng people’s hospital, Liaocheng, Shandong, 252000, China; Department of Pulmonology, The Affiliated Hospital of Shandong University of TCM, Jinan, Shandong, 250000, China; Department of Respiratory Medicine, The First Affiliated Hospital of Shandong First Medical University, Jinan, Shandong, 250000, China; Department of Central laboratory, Liaocheng people’s hospital, Liaocheng, Shandong, 252000, China

**Keywords:** COVID-19, Cluster, Epidemiology, Generation, Clinical, Characteristics

## Abstract

**Objective:** To analyze the epidemiological and intergenerational clinical characteristics of COVID-19 patients associated with cluster, so as to understand the rules of the patients associated with cluster of this outbreak and provide help for the prevention and control of COVID-19.

**Methods:** All close contacts of the patient were screened since the first supermarket employee with COVID-19 was identified. A retrospective analysis was made on the epidemiological and clinical characteristics of the confirmed cases admitted to the designated hospitals for centralized treatment. The patients were divided into two groups according to the first generation (supermarket employees, group A) and the second or third generation (family members or friends of supermarket employees, group B), and the similarities and differences between the two groups were compared.

**Results:** A total of 24 COVID-19 patients were diagnosed, with an average age of 48 ±1.73 years. The mean duration from onset to release form quarantine was 21.04± 6.77 days, and the onset time was concentrated in 5-11 days after the first patient was diagnosed. Among all the patients, 23 patients were moderate, among which 7 patients (29.17%) were asymptomatic. Symptoms of symptomatic patients were cough (75.00%), low fever (62.50%), shortness of breath (41.67%), sore throat (25.00%), gastrointestinal symptoms (25.00%), fatigue (20.83%), etc. Biochemical examination on admission showed that the white blood cell count < 4.0×109/L (29.17%) and the lymphocyte count <1.1×109/L (58.33%). The lymphocyte count of 50.00% of the patients was ≤ 0.6 × 109/L. On admission, chest CT showed pneumonia (100%) with bilateral infiltration (75.00%). Treatment: antiviral drug (100%), Chinese medicine (100%), common oxygen therapy (45.83%). There were 11 cases in group A (first generation, 11 cases) and 13 cases in group B (second generation, 11 cases; third generation, 2 cases). In group B, there were more males, from onset to admission later, more patients had underlying diseases, and more patients were treated with albumin (P<0.05). However, there was no statistical difference between the two groups in other clinical indicators, including the duration from onset to release form quarantine(P>0.05). There was no improvement in granulocyte count in all patients, as well as in groups A and B, between admission and release from quarantine(P>0.05).

**Conclusion:** The clinical characteristics of COVID-19 patients associated with cluster were similar to those of other COVID-19 patients, but there were some special features. The severity of the disease was similar and there was intergenerational spread. There was no difference in clinical characteristics between generations. Asymptomatic infections occurred in a proportion of patients and could cause spread.

## Background

COVID-19 is widespread in the world, and there have been many summaries on the clinical characteristics ^[1,2,3,4]^, and many research articles have been published on the epidemiological characteristics of COVID-19^[5,6,7,8]^.

Liaocheng City, in the Middle East of China, far from Hubei province and Wuhan city, had a total of 38 confirmed COVID-19 patients in this outbreak. Among these patients, there were 24 COVID-19 patients associated with supermarket cluster, accounting for 63.2% (24/38) of all patients, which was one of the main causes of COVID-19 in non-outbreak areas. Analysis of the epidemiological and clinical characteristics of these patients is of great significance for the prevention and treatment of COVID-19.

## Methods

The monitoring, treatment, release from quarantine and severity of all cases were determined in accordance with the protocol of diagnosis and treatment of novel coronavirus pneumonia issued by National Health Commission of the People’s Republic of China and National Administration of Traditional Chinese Medicine ^[9,10,11]^. A confirmed case was defined as a positive result to real-time reverse-transcriptase polymerase-chain-reaction (RT-PCR) assay for nasal and pharyngeal swab specimens ^[12]^.

Screening and diagnosis: after the first supermarket employee was diagnosed with COVID-19 due to illness, all of the patient’s close contacts were then screened, those who had been to the supermarket a week before the time of the patient’s last visit. Subjects screened with or without respiratory symptoms or other symptoms were routinely given RT-PCR assay for nasal and pharyngeal swab specimens, followed by routine screening of family and friends with positive confirmed cases, the method was the same as above. All these confirmed patients were given chest CT examination and received centralized monitoring and treatment in designated hospitals.

The criteria for all cases to be released from quarantine were as recommended in the above protocol,except for the obvious improvement of clinical manifestations and imaging of acute exudative lesions, the focus was on RT-PCR detection of nucleic acid. After 5 days in hospital, every 2 days test, to nucleic acid test negative case, instead of every 24 hours test, three consecutive tests were negative, they can be released from quarantine. During the dynamic test, if nucleic acid negative cases were once again positive or suspicious, the above steps were restarted. Some cases were kept in hospital for 14 days after they were released from quarantine.

The duration from onset to admission of asymptomatic screening confirmed infected patients was calculated as 1 day. The length of hospital stay was the time from admission to the release form quarantine, and the time still observed in the hospital after release form quarantine was not included in the length of hospital stay. The sum of these two periods was defined as the time from onset to release form quarantine.

This study retrospectively analyzed the general characteristics, epidemiological history, chronic underlying diseases, clinical symptoms and complications, chest CT, biochemical monitoring, severity assessment, treatment and outcome of 24 COVID-19 patients associated with supermarket cluster. In addition, the patients were divided into two groups according to the first generation (supermarket employees, group A) and the second or third generation (family members or friends of supermarket employees, group B), and the clinical characteristics of the two groups were compared.

## Statistical analysis

Continuous variables were expressed as the means and standard deviations or medians and interquartile ranges (IQR) as appropriate. Categorical variables were summarized as the counts and percentages in each category. T test or Wilcoxon rank-sum tests were applied to continuous variables. Chi-square test or Fisher’s exact tests were used for categorical variables. All analyses were conducted with SPSS software version 23.0.

## Results

During the outbreak, about 8,000 people were screened and a total of 24 COVID 19 patients were confirmed, including 11 supermarket employees and 13 employees’ family members or friends (10 family members and 3 friends). No supermarket customers were diagnosed. All the cases associated with the supermarket cluster had no history of sojourn in the epidemic area, among which the secondary cases due to close contact had a history of close contact with the confirmed supermarket staff. 7 of the patients were asymptomatic, 6 of them were supermarket employees, and 3 of them caused 6 family members or friends to have symptoms. The onset time of symptomatic patients was concentrated between 5 and 11 days after the onset of symptoms in the first confirmed patient (figure I). Since it was difficult to determine the source of infection of the confirmed supermarket employees, which was identified as the common exposure, 3 generations of cases were generated in the clusters, including 11 cases in the first generation (11/24, 45.8%), 11 cases in the second generation (11/24, 45.8%), and 2 cases in the third generation (2/24, 8.3%) (figure II). The incubation period of these patients was also difficult to determine.

**Figure I:**
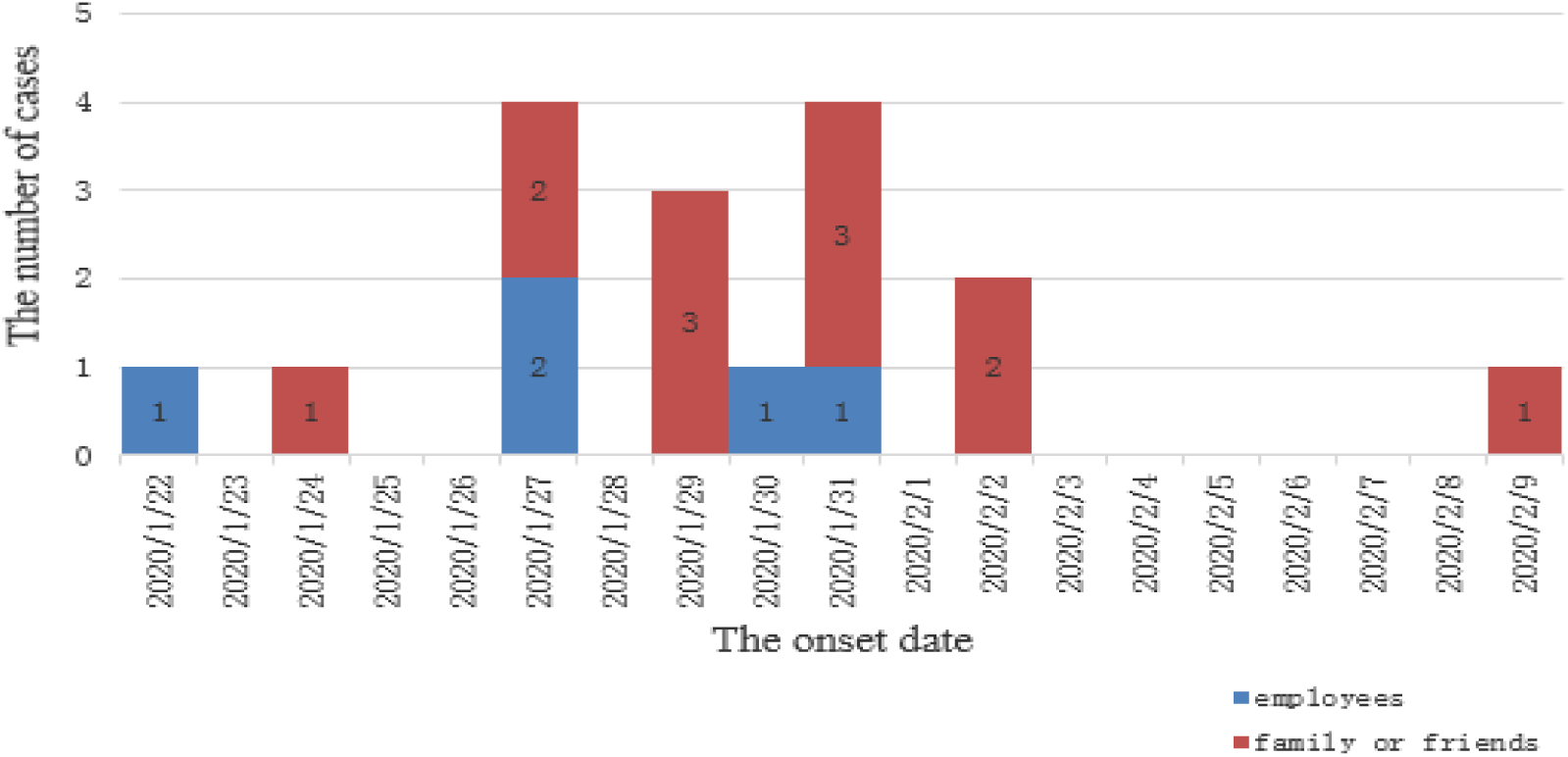
Time distribution of confirmed symptomatic COVID-19 patients associated with supermarket cluster

**Figure II:**
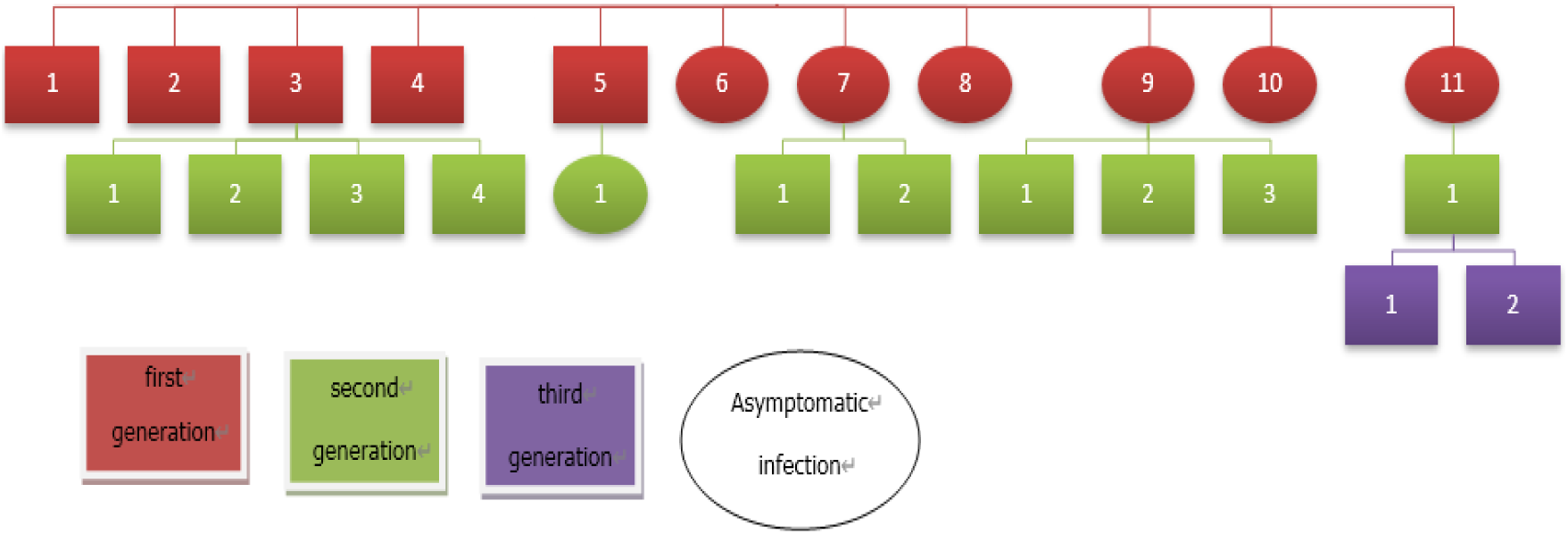
Transmission chain and intergenerational relationship

The first case with an unknown epidemiological history was diagnosed with fever and sore throat. The time from onset to release from quarantine was the longest of these patients, 34 days. The shortest was 8 days and the patient was diagnosed by screening. The third confirmed patient suffered from fever, and his parent, spouse and two children were all diagnosed as COVID-19 with fever, and three of them suffered from fever on the same day. However, the time from onset to release from quarantine was different among the five people, ranging from 17 days to 30 days.

According to the Chinese diagnosis and treatment protocol of COVID-19^[9,10,11]^, the patients were divided into mild, moderate, severe and critical according to the severity of the disease. The difference between mild and moderate patients was the presence of pneumonia, and the patients with respiratory dysfunction or multiple organ dysfunction syndrome (MODS) were severe or critical. 23 of the 24 patients in this group were moderate. However, according to the pneumonia severity index (PSI) score, 83.3%(16+4/24) of the patients were mild patients of grade I and II, 4 patients of grade III and IV did not develop severe or critical disease, and 1 patient of grade II, aged, with a history of diabetes, developed severe disease due to ARDS on the 20 days after the diagnosis of COVID 19. Admission symptoms, in addition to asymptomatic infection patients, the most common symptom was cough, followed by fever, and were low fever, other symptoms were successively shortness of breath, sore throat, gastrointestinal symptoms, fatigue. On the biochemical examination of admission, in addition to the white blood cell and lymphocyte counts, there were some changes beyond the normal range, but the absolute value of the change was not significant. It was important to note that all patients were admitted with chest CT scans showing pneumonia, often with bilateral infiltrations, including 7 asymptomatic patients. In terms of treatment during hospitalization, all patients were given 2-3 antiviral drugs, and some patients were given antibiotics, hormones and immunomodulators. Except for 1 case of severe COVID-19 was treated with High-Flow Nasal Cannula(HFNC), all the others were normal oxygen therapy. The application proportion of Traditional Chinese Medicine (TCM) was 100%, including Chinese medicine preparation, acupuncture, moxibustion, etc. (Table I, Table II).

**Table 1.** Clinical Characteristics of the Study Patients on Admission

| Characteristics | All Patients<br>(n=24) | Group |  | P |
| --- | --- | --- | --- | --- |
|  |  | A<br>(n=11) | B<br>(n=13) |  |
| Age (y) | 48±1.73 | 42.05±9.46 | 53±2.09 | 0.11 |
| Gender |  |  |  |  |
| Male (%) | 9/24(37.50) | 1/11(9.09) | 8/13(61.54) | 0.013 |
| Female (%) | 15/24(62.50) | 10/11(90.91) | 5/13(38.46) |  |
| Duration from onset to hospitalization(d) | 5.5 (7) | 3 (5) | 6 (4) | 0.011 |
| PSI | 0 (57.25) | 0 (44) | 45 (72.5) | 0.252 |
| I≤50 (%) | 16/24(66.67) | 9/11(81.82) | 7/13(53.85) |  |
| II 51-70 (%) | 4/24(16.67) | 1/11(9.09) | 3/13(23.08) |  |
| III 71-90 (%) | 3/24(12.50) | 1/11(9.09) | 2/13(15.38) |  |
| IV91-131 (%) | 1/24(4.17) | 0/11(0) | 1/13(7.69) |  |
| V>130 (%) | 0/37(0) | 0/19(0) | 0/18(0) |  |
| Underlying disease |  |  |  |  |
| Any (%) | 6/24(25.00) | 0/11(0) | 6/13(46.15) | 0.016 |
| Hypertension (%) | 2/24(8.33) | 0/11(0) | 2/13(15.38) | 0.482 |
| Coronary heart disease (%) | 2/24(8.33) | 0/11(0) | 2/13(15.38) | 0.482 |
| Diabetes (%) | 1/24(4.17) | 0/11(0) | 1/13(7.69) | 1 |
| Cirrhosis, Liver cancer (%) | 1/24(4.17) | 0/11(0) | 1/13(7.69) | 1 |
| Symptoms |  |  |  |  |
| Asymptomatic (%) | 7/24(29.17) | 6/11(54.55) | 1/13(7.69) | 0.023 |
| Fever (%) | 15/24(62.50) | 5/11(45.45) | 10/13(76.92) | 0.206 |
| Fatigue (%) | 5/24(20.83) | 1/11(9.09) | 4/13(30.77) | 0.327 |
| Headache (%) | 1/24(4.17) | 1/11(9.09) | 0/13(0) | 0.458 |
| Nasal congestion (%) | 2/24(8.33) | 0/11(0) | 2/13(15.38) | 0.482 |
| Sore throat (%) | 6/24(25.00) | 2/11(18.18) | 4/13(30.77) | 0.649 |
| Cough (%) | 18/24(75.00) | 8/11(72.73) | 10/13(76.92) | 1 |
| Hemoptysis (%) | 1/24(4.17) | 0/11(0) | 1/13(7.69) | 1 |
| Shortness of breath (%) | 10/24(41.67) | 3/11(27.27) | 7/13(53.85) | 0.240 |
| Vomiting or diarrhea (%) | 6/24(25.00) | 3/11(27.27) | 3/13(23.08) | 1 |
| Pain in a muscle or joint (%) | 1/24(4.17) | 0/11(0) | 1/13(7.69) | 1 |
| Laboratory findings |  |  |  |  |
| WBC<4.0×10<sup>9</sup>/L (%) | 7/24(29.17) | 3/11(27.27) | 4/13(30.77) | 1 |
| L <1.1×10<sup>9</sup>/L (%) | 14/24(58.33) | 6/11(54.55) | 8/13(61.54) | 1 |
| L≤0.6×10<sup>9</sup>/L (%) | 7/14(50.00) | 3/6(50.0) | 4/8(50.00) | 1 |
| SAA>10mg/L (%) | 15/24(62.50) | 5/11(45.45) | 10/13(76.92) | 0.206 |
| CRP>5mg/L (%) | 12/24(50.00) | 3/11(27.27) | 9/13(69.23) | 0.1 |
| PCT>0.5ng/ml (%) | 2/24(8.33) | 2/11(18.18) | 0/13(0) | 0.199 |
| LDH>250u/L (%) | 5/24(20.83) | 1/11(9.09) | 4/13(30.77) | 0.327 |
| Alb <40g/L (%) | 17/24(70.83) | 8/11(72.73) | 9/13(69.23) | 1 |
| AST>40u/L (%) | 2/24(8.33) | 1/11(9.09) | 1/13(7.69) | 1 |
| ALT>40u/L (%) | 1/24(4.17) | 1/11(9.09) | 0/13(0) | 0.458 |
| TBL>17.1ummol/L (%) | 9/24(37.50) | 3/11(27.27) | 6/13(46.15) | 0.423 |
| D -dimer >0.5mg/L (%) | 6/24(25.00) | 3/11(27.27) | 3/13(23.08) | 1 |
| Fib>4g/L ( % ) | 6/24(25.00) | 3/11(27.27) | 3/13(23.08) | 1 |
| ESR≥20mm/h ( % ) | 18/24(75.00) | 8/11(72.73) | 10/13(76.92) | 1 |
| Chest CT |  |  |  |  |
| Pneumonia ( % ) | 24/24(100) | 11/11(100) | 13/13(100) | -- |
| Bilateral infiltration ( % ) | 18/24(75.00) | 7/11(63.64) | 11/13(84.62) | 0.357 |
| Unilateral infiltration ( % ) | 6/24(25.00) | 4/11(36.36) | 2/13(15.38) |  |

PSI: Pneumonia Severity Index; WBC: White Blood Cells; L: Lymphocyte; SAA: Serum Amyloid A; CRP: C-Reactive Protein; PCT: Procalcitonin; LDH: Lactate Dehydrogenase; Alb: Albumin; AST: Aspartate Aminotransferase; ALT: Alanine Aminotransferase; TBL: Total Bilirubin; Fib: Fibrinogen; ESR: Erythrocyte Sedimentation Rate.

**Table II.** Severity Classification of Patients and Complications and Treatment Measures before Release form Quarantine

| Characteristics | All Patients<br>(n=24) | Group |  | P |
| --- | --- | --- | --- | --- |
|  |  | A<br>(n=11) | B<br>(n=13) |  |
| Severity |  |  |  | 0.776 |
| Mild ( % ) | 0/24(0) | 0/11(0) | 0/13(0) |  |
| Moderate ( % ) | 23/24(95.83) | 11/11(100) | 12/13(92.31) |  |
| Severe ( % ) | 1/24(4.17) | 0/11(0) | 1/13(7.69) |  |
| Critical ( % ) | 0/24(0) | 0/11(0) | 0/13(0) |  |
| Complications |  |  |  |  |
| ARDS ( % ) | 1/24 (4.17) | 0/11(0) | 1/13(7.69) | 1 |
| Treatment |  |  |  |  |
| Antibiotics | 17/24 (70.83) | 9/11 (81.82) | 8/13 (61.54) | 0.386 |
| Antifungal drugs | 1/24(4.17) | 0/11(0) | 1/13(7.69) | 1 |
| Antiviral drugs ( % ) | 24/24(100) | 11/11(100) | 13/13(100) | -- |
| Glucocorticoids ( % ) | 5/24(20.83) | 2/11(18.18) | 3/13(23.08) | 1 |
| Albumin ( % ) | 11/24(45.83) | 2/11(18.18) | 9/13(69.23) | 0.019 |
| Immunoglobulin ( % ) | 6/24(25.00) | 2/11(18.18) | 4/13(30.77) | 0.649 |
| Thymosin ( % ) | 15/24(62.50) | 7/11(63.64) | 8/13(61.54) | 1 |
| Oxygen therapy ( % ) | 11/24(45.83) | 4/11(36.36) | 7/13(53.85) | 0.444 |
| Common ( % ) | 10/11(90.91) | 4/4(100) | 6/7(85.71) | 1 |
| HFNC ( % ) | 1/11(9.09) | 0/4(0) | 1/7(14.29) |  |
| TCM ( % ) | 24/24(100) | 11/11(100) | 13/13(100) | -- |

HFNC: High-Flow Nasal Cannula; PE: Plasma Exchange; TCM: Traditional Chinese Medicine.

Compared with group B, there were more men in group B, from onset to admission later, more underlying diseases, and more cases were treated with albumin (P<0.05). There were no statistical differences in other indicators (Table I, Table II, and Table III).

**Table III.** Clinical Outcomes of Patients

| Outcomes | All Patients<br>(n=24) | Group |  |  |
| --- | --- | --- | --- | --- |
|  |  | A<br>(n=11) | B<br>(n=13) | P |
| Hospitalization duration (d) | 16±6.15 | 16±6.66 | 16±5.96 | 1 |
| Duration from onset to release from quarantine (d) | 21.04±6.77 | 19.09±7.92 | 22.69±5.41 | 0.201 |
| Blood findings at the time of release from quarantine |  |  |  |  |
| WBC <4.0×10<sup>9</sup>/L (%) | 3/24(12.50) | 1/11(9.09) | 2/13(15.38) | 1 |
| L <1.1×10<sup>9</sup>/L (%) | 14/24(58.33) | 7/11(63.64) | 7/13(53.85) | 0.697 |
| L ≤0.6×10<sup>9</sup>/L (%) | 4/14(28.57) | 1/7(14.29) | 3/7(42.86) | 0.559 |
| SAA>10mg/L (%) | 4/24(16.67) | 1/11(9.09) | 3/13(23.08) | 0.596 |

There was no significant change in the granulocyte count of all patients or of patients in groups A and B on admission and release from quarantine (Table IV).

**Table IV.** Granulocyte Counts on Admission and Release from Quarantine

| Granulocyte counts | Group |  |  |  |  |  |  |  |  |
| --- | --- | --- | --- | --- | --- | --- | --- | --- | --- |
|  | All patients (n=24) |  |  | A<br>(n=11) |  |  | B<br>(n=13) |  |  |
|  | Admission | Release | P | Admission | Release | P | Admission | Release | P |
| WBC (×10<sup>9</sup>/L) | 4.88±2.57 | 5.21±1.55 | 0.89 | 5.77±2.91 | 5.25±1.46 | 0.598 | 4.87±1.57 | 5.18±1.68 | 0.641 |
| L (×10<sup>9</sup>/L) | 0.86 (0.75) | 0.86 (0.6) | 0.672 | 0.99±0.40 | 0.99±0.33 | 0.986 | 1.0±0.66 | 1.02±0.45 | 0.945 |
| WB < 4.0×10<sup>9</sup>/L (%) | 7/24(29.17) | 3/24(12.50) | 0.286 | 3/11(27.27) | 1/11(9.09) | 0.586 | 4/13(30.77) | 2/13(15.38) | 0.645 |
| L <1.1×10<sup>9</sup>/L (%) | 14/24(58.33) | 14/24(58.33) | 1 | 6/11(54.55) | 7/11(63.64) | 1 | 8/13(61.54) | 7/13(53.85) | 1 |
| L ≤0.6×10<sup>9</sup>/L (%) | 7/14(50.00) | 4/14(28.57) | 0.44 | 3/6(50.00) | 1/7(14.29) | 0.266 | 4/8(50.00) | 3/7(42.86) | 1 |

## Discuss

The control of COVID-19 associated with cluster was one of the key links in the prevention and control of COVID-19 outbreak ^[13,14]^. After the diagnosis of the first case, all colleagues and customers within a period of time were quickly screened, and close contacts of confirmed patients were screened again. These measures could be timely detection and control the source of infection. This was the most important part to effectively control the further spread of this outbreak.

The onset time of this group of patients was mainly about a week after the first patient was diagnosed. There were three generations of intergenerational relationship. However, most of them had been limited to two generations. Other studies on cluster cases of COVID-19 were mostly due to family cluster, and the number of cases was also small _[15,16,17]_.

Asymptomatic infections were attracting more and more attention ^[16,18,19]^, not only because these patients were not rare, but also had obvious infectivity, and this study also showed the same results.

Because the patients in this study were mainly diagnosed through screening, the patients did not develop serious clinical manifestations during their hospitalization. Although several patients had slightly higher PSI scores, this was due to underlying disease and age. They did not meet the diagnostic criteria for severe or critical type. A patient with PSI score of grade II developed severe during hospitalization because the change occurred 20 days after the diagnosis of COVID 19 and was not directly related to novel coronavirus. Therefore, these COVID-19 patients associated with supermarket cluster were all of the moderate type, the severity of the disease did not differ significantly, it was not clear whether this was a feature of patients associated with cluster.

In terms of admission symptoms, there was no significant difference between the study patients in this group and other studies on the clinical characteristics of COVID 19 _[1,2,3,4]_.

It was worth noting that, whether the patient had clinical symptoms or not, all the patients in this group had chest CT manifestations of pneumonia, and most of them were bilateral infiltration, which was different from other respiratory virus infections _[20,21]_.

As for the treatment regimen, due to the particularity of the outbreak, according to the above recommendations on the treatment protocol of COVID-19, although there was no specific antiviral drugs at present, many studies had suggested the benefits of early use of antivirals ^[22,23]^. The patients in this group were also given at least two antiviral drugs. There was no sufficient evidence for the application of antibiotics, hormones and immune modulators in some patients. All patients were routinely treated with traditional Chinese medicine (TMC), and many studies had demonstrated its important role in inhibiting coronavirus^[24,25,26]^.

The most important indicator for release from quarantine of the patients in the group was the continuous negative result of novel coronavirus nucleic acid test. Therefore, the duration from onset to release form quarantine reflected the time it took the patient to release the virus from the respiratory tract. There were intergenerational differences in the duration from onset to admission and in the underlying disease, however, in the end, all the patients in the two groups were released from quarantine, and the duration from onset to release from quarantine was not statistically significant, which may be due to the small number of patients in this group and the patient’s condition is not serious, etc. In addition, the first supermarket employee diagnosed with symptoms had the longest duration from onset to release from quarantine. The third supermarket employee diagnosed with symptoms caused familial clustering COVID-19, with the same symptoms but significantly different duration of release from quarantine. These also suggested that the cause of respiratory novel coronavirus positive duration was complex ^[27,28]^.

In general, the basic clinical characteristics of the patients in this study group were similar to those of other COVID 19 patients ^[1,2]^. Although some clinical characteristics were different between the first generation of patients and the second and third generation of patients, this was related to the timeliness of screening and the characteristics of the population. There was no statistical difference in the clinical manifestations between the two groups after the onset, even including the duration from onset to release from quarantine.

The reduction in granulocyte count caused by novel coronavirus has been a concern ^[27,29]^, and the patients in the group showed the same characteristics, especially lymphocytes, and about half of the patients had lymphocyte counts less than 0.6×109/L, which seriously affected the immune function of the body. Moreover, the white blood cell and lymphocyte counts at the time of release from quarantine did not improve significantly compared with those on admission, both in all patients and in groups A and B, which required continuous follow-up observational studies.

The patients studied in this group were cases associated with cluster, and the number of patients was not large and there were regional limitations. Therefore, these cases did not represent the characteristics of patients with sporadic COVID-19 in a wider area.

## Conclusion

1. The clinical characteristics of COVID-19 patients associated with cluster were similar to those of other COVID-19 patients, but there were some special features.
2. The severity of the disease was similar and there was intergenerational spread. Although the basic characteristics before the onset of the disease were different between generations, the clinical manifestations after the onset were not different.
3. Asymptomatic infection of COVID-19 must cause enough attention, not only the proportion was higher, but also spread.

## Data Availability

With the permission of the corresponding author, we can provide participant data,statistical analysis.

---

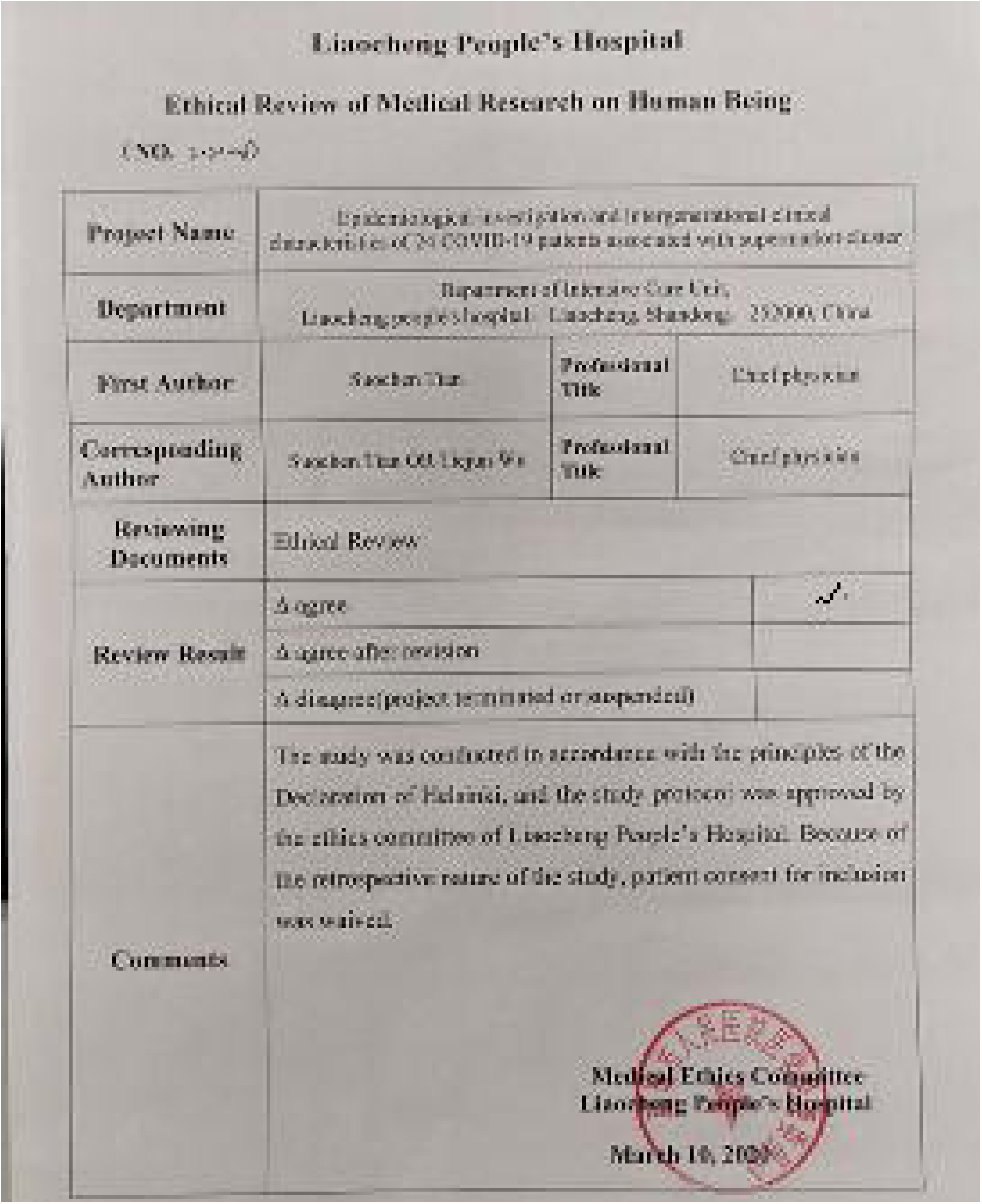

## Notes

**Declaration of Interest** All authors have no conflicts of interest to declare

### Competing Interest Statement

The authors have declared no competing interest.

### Funding Statement

We haven't received any external funding.

